# Co-designing a digital health app to manage pain in young children with cancer: report from the generative design phase of intervention development

**DOI:** 10.1101/2022.11.23.22282681

**Authors:** Lindsay A. Jibb, Surabhi Sivaratnam, Elham Hashemi, Jennifer N. Stinson, Paul C. Nathan, Julie Chartrand, Nicole M. Alberts, Tatenda Masama, Hannah G. Pease, Lessley B. Torres, Haydee G. Cortes, Mallory Zworth, Susan Kuczynski, Michelle A. Fortier

## Abstract

Pain is one of the most prevalent and burdensome pediatric cancer symptoms for young children and their families. A significant proportion of pain episodes are experienced in environments where management options are limited, including at home, and digital innovations such as apps may have positive impacts on pain outcomes for young children in these environments. Our overall aim is to co-design such an app and the objective of this study was to explore the perceptions of children’s parents about app utility, needed system features, and challenges. We recruited parents of young children with cancer and multidisciplinary pediatric oncology clinicians from two pediatric cancer care centers to participate in audio-recorded, semi-structured co-design interviews. We conducted interviews until data saturation was reached. Audio-recordings were then transcribed, coded, and analyzed using thematic analysis. Forty-two participants took part in the process. Participants endorsed the concept of an app as a useful, safe, and convenient way to engage caregivers in managing their young child’s pain. The value of the app related to its capacity to provide real-time, multimodal informational and procedural pain support to parents, while also reducing the emotional burden of pain care. Recommendations for intervention design included accessibility-focused features, comprehensive symptom tracking, and embedded scientific- and clinically-sound symptom assessments and management advice. Predicted challenges associated with digital pain management related to potential burden of use for parents and clinicians. The insights gathered will inform the design principles of our future childhood cancer pain digital research.

**AUTHOR SUMMARY:** The lack of meaningful involvement of end-users in intervention development has been a key contributor to difficulties in effectively translating research findings into cancer practice and policy. There is a risk that without the active engagement of children with cancer and their families in designing digital health innovations, researchers and clinicians will fall victim to an unfortunate cycle of producing underutilized evidence—resulting in a limited impact on patient outcomes. Pain is a particular problem for young children with cancer and real-time digital health interventions may be solutions for accessible, effective, and scalable cancer pain management. We are using an established end user-centered co-design process to engage parents and pediatric oncology clinicians in the development of a cancer pain management app. Our work here summarizes the generative co-design phase of this process and the perceptions of parents and clinicians related to app usefulness and needed system features.

## INTRODUCTION

Pain is one of the most prevalent and burdensome pediatric cancer symptoms for children and their families despite the existence of evidence-based treatment guidelines (1,2). The negative consequences of childhood cancer pain are many and include reduced child health-related quality of life, increased child and family distress, chronic pain in survivorship and the potential for significant financial costs to healthcare systems and families (3–6). Further, shifts to increasingly outpatient-based cancer care mean that children with cancer are experiencing pain in environments, such as home or school, where treatment options are limited (7).

Young children with cancer, including toddlers, preschoolers, and school-aged children, are particularly vulnerable to undermanaged pain due to their limited ability for pain self-report and their reliance on caregivers for pain management and treatment. Current research shows that digital innovations such as real-time smartphone-based pain management support apps may have positive impacts on pain outcomes in adolescents with cancer (5,8–10), but no investigations have been conducted into such tools for managing pain in young children with cancer, especially outside the hospital setting.

To increase the likelihood of successful implementation of digital health interventions within pediatric cancer care, it is vital that the key stakeholders, including parents, other family caregivers and clinicians, are meaningfully integrated in the processes of intervention design, development, and evaluation. Evidence shows that eliciting stakeholder perspectives, including that of parents, on intervention development supports the identification of barriers to use, results in interventions with enhanced perceived effectiveness, and facilitates the successful delivery of digital health interventions (11–13).

The current study is part of a phased approach to the development and evaluation of a digital health app for the parent-led management of young children’s cancer pain. Our process employed a co-design framework, which has been successfully used to develop pediatric health innovations (14). This study describes the generative design phase of innovation development wherein the healthcare needs of participants, including latent needs that participants may not be aware of, are explored and revealed. Our future co-design efforts will build on these results during the software development and co-evaluation of a high-fidelity app prototype with family caregivers.

### Objective

Given the issue of undermanaged pain in young children with cancer and the critical need to elicit stakeholders input early in the process of successful digital health intervention design, the objective of this study was to explore the perceptions of children’s parents and clinicians as they pertain to a the co-design of a parent-led real-time cancer pain management app, including possible utility, recommendations for needed system features, and potential challenges to implementation.

## RESULTS

### Study sample

A total of 42 participants—21 parents and 21 clinicians—were interviewed between December 2019 and September 2020. All eligible participants who were approached agreed to participate. The mean (range) total interview times were 33 (14-67) minutes and 33 (22-45) minutes for parents and clinicians, respectively. Participant characteristics are shown in **Table 1**. Most parents were within the 30-39 years of age category (n=13, 62%) and mothers (n=17, 81%). The mean parent-reported PPEP scores were 33 (SD=10), indicating moderately low child pain misconceptions. Clinicians were most often registered nurses (n=9, 43%) and had a mean average 10 years of clinical experience.

**Table 1.**
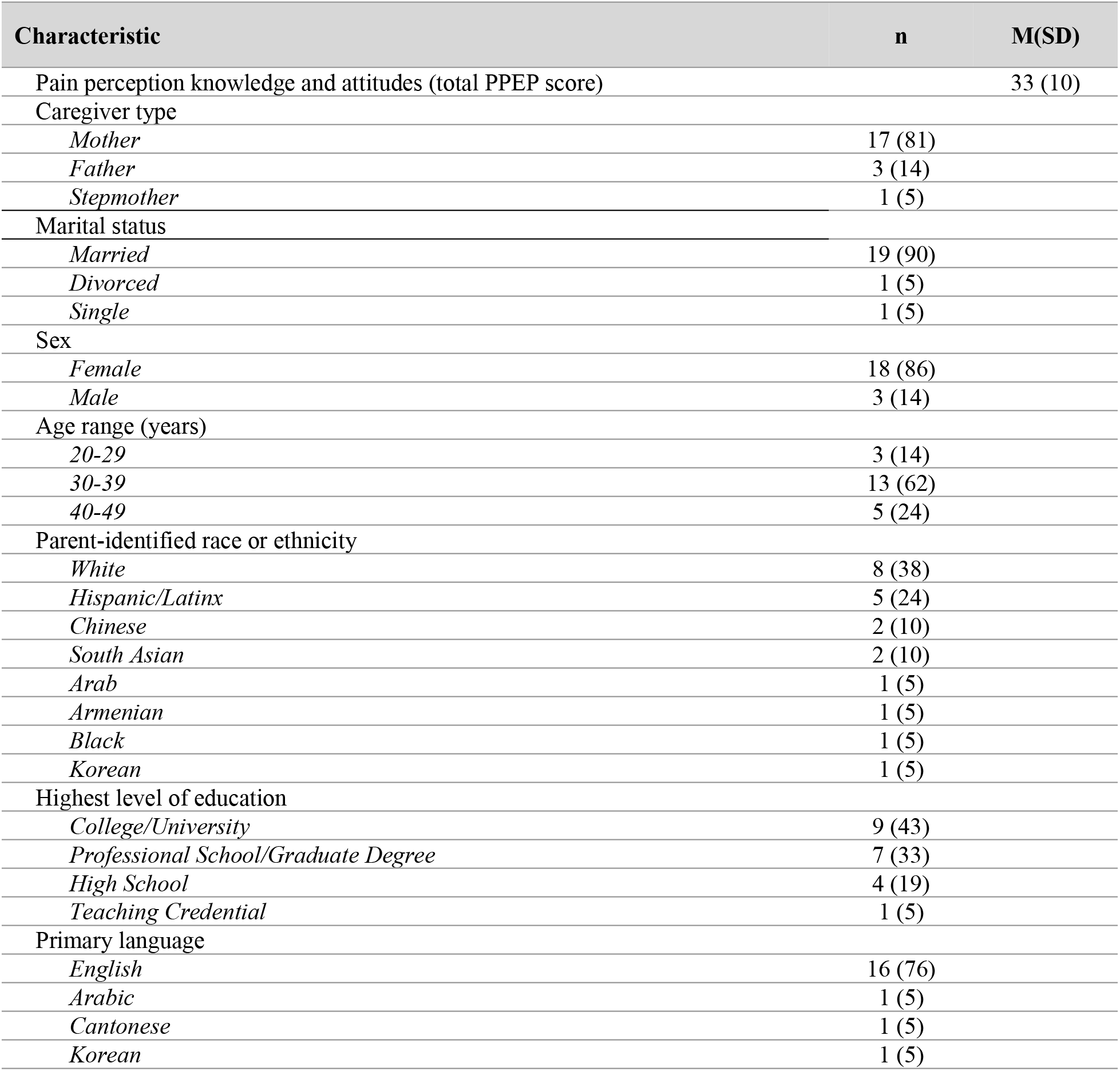
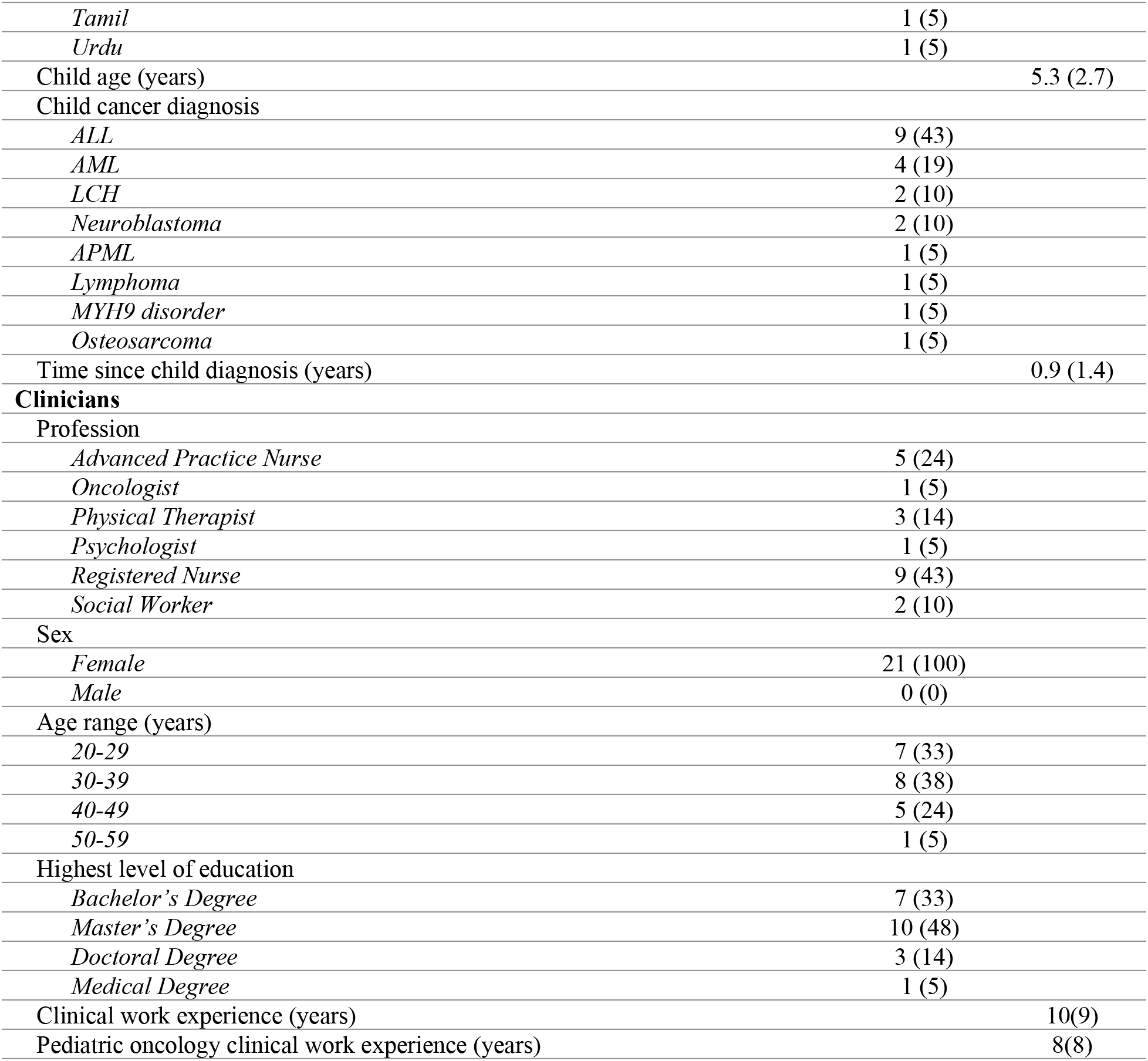
Participant characteristics

| Characteristic | n | M(SD) |
| --- | --- | --- |
| Pain perception knowledge and attitudes (total PPEP score) |  | 33 (10) |
| Caregiver type |  |  |
| *Mother* | 17 (81) |  |
| *Father* | 3 (14) |  |
| *Stepmother* | 1 (5) |  |
| Marital status |  |  |
| *Married* | 19 (90) |  |
| *Divorced* | 1 (5) |  |
| *Single* | 1 (5) |  |
| Sex |  |  |
| *Female* | 18 (86) |  |
| *Male* | 3 (14) |  |
| Age range (years) |  |  |
| 20-29 | 3 (14) |  |
| 30-39 | 13 (62) |  |
| 40-49 | 5 (24) |  |
| Parent-identified race or ethnicity |  |  |
| *White* | 8 (38) |  |
| *Hispanic/Latinx* | 5 (24) |  |
| *Chinese* | 2 (10) |  |
| *South Asian* | 2 (10) |  |
| *Arab* | 1 (5) |  |
| *Armenian* | 1 (5) |  |
| *Black* | 1 (5) |  |
| *Korean* | 1 (5) |  |
| Highest level of education |  |  |
| *College/University* | 9 (43) |  |
| *Professional School/Graduate Degree* | 7 (33) |  |
| *High School* | 4 (19) |  |
| *Teaching Credential* | 1 (5) |  |
| Primary language |  |  |
| *English* | 16 (76) |  |
| *Arabic* | 1 (5) |  |
| *Cantonese* | 1 (5) |  |
| *Korean* | 1 (5) |  |
| *Tamil* | 1 (5) |  |
| *Urdu* | 1 (5) |  |
| Child age (years) |  | 5.3 (2.7) |
| Child cancer diagnosis |  |  |
| *ALL* | 9 (43) |  |
| *AML* | 4 (19) |  |
| *LCH* | 2 (10) |  |
| *Neuroblastoma* | 2 (10) |  |
| *APML* | 1 (5) |  |
| *Lymphoma* | 1 (5) |  |
| *MYH9 disorder* | 1 (5) |  |
| *Osteosarcoma* | 1 (5) |  |
| Time since child diagnosis (years) |  | 0.9 (1.4) |
| **Clinicians** |  |  |
| Profession |  |  |
| *Advanced Practice Nurse* | 5 (24) |  |
| *Oncologist* | 1 (5) |  |
| *Physical Therapist* | 3 (14) |  |
| *Psychologist* | 1 (5) |  |
| *Registered Nurse* | 9 (43) |  |
| *Social Worker* | 2 (10) |  |
| Sex |  |  |
| *Female* | 21 (100) |  |
| *Male* | 0 (0) |  |
| Age range (years) |  |  |
| *20-29* | 7 (33) |  |
| *30-39* | 8 (38) |  |
| *40-49* | 5 (24) |  |
| *50-59* | 1 (5) |  |
| Highest level of education |  |  |
| *Bachelor's Degree* | 7 (33) |  |
| *Master's Degree* | 10 (48) |  |
| *Doctoral Degree* | 3 (14) |  |
| *Medical Degree* | 1 (5) |  |
| Clinical work experience (years) |  | 10(9) |
| Pediatric oncology clinical work experience (years) |  | 8(8) |

### Thematic analysis

We organized our data into four major themes to describe parent and clinician perceptions of the utility of digital cancer pain intervention, recommendations for intervention components, and potential implementation challenges. **Table 2** shows themes, subthemes, and example narrative quotes.

**Table 2.**
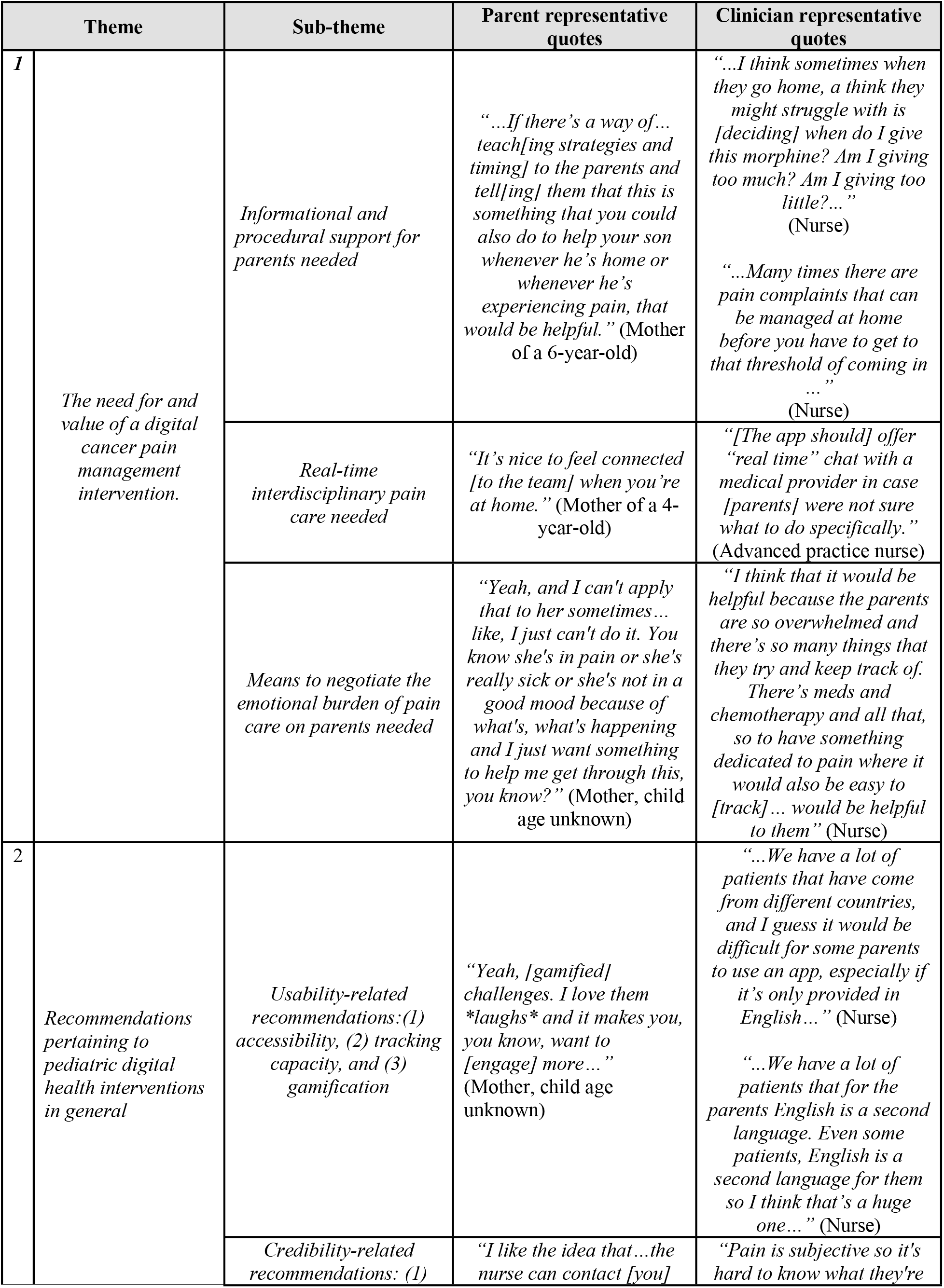
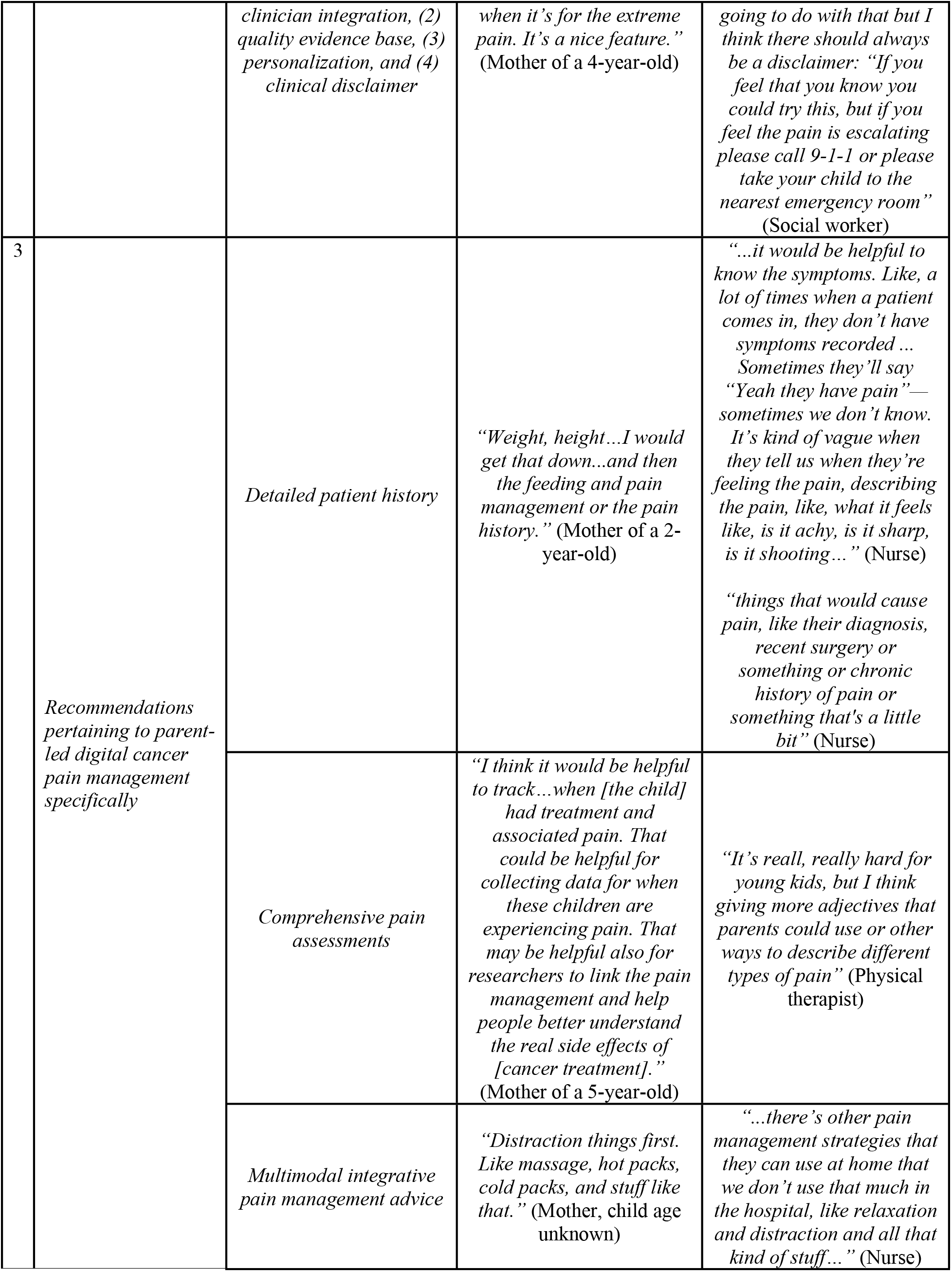
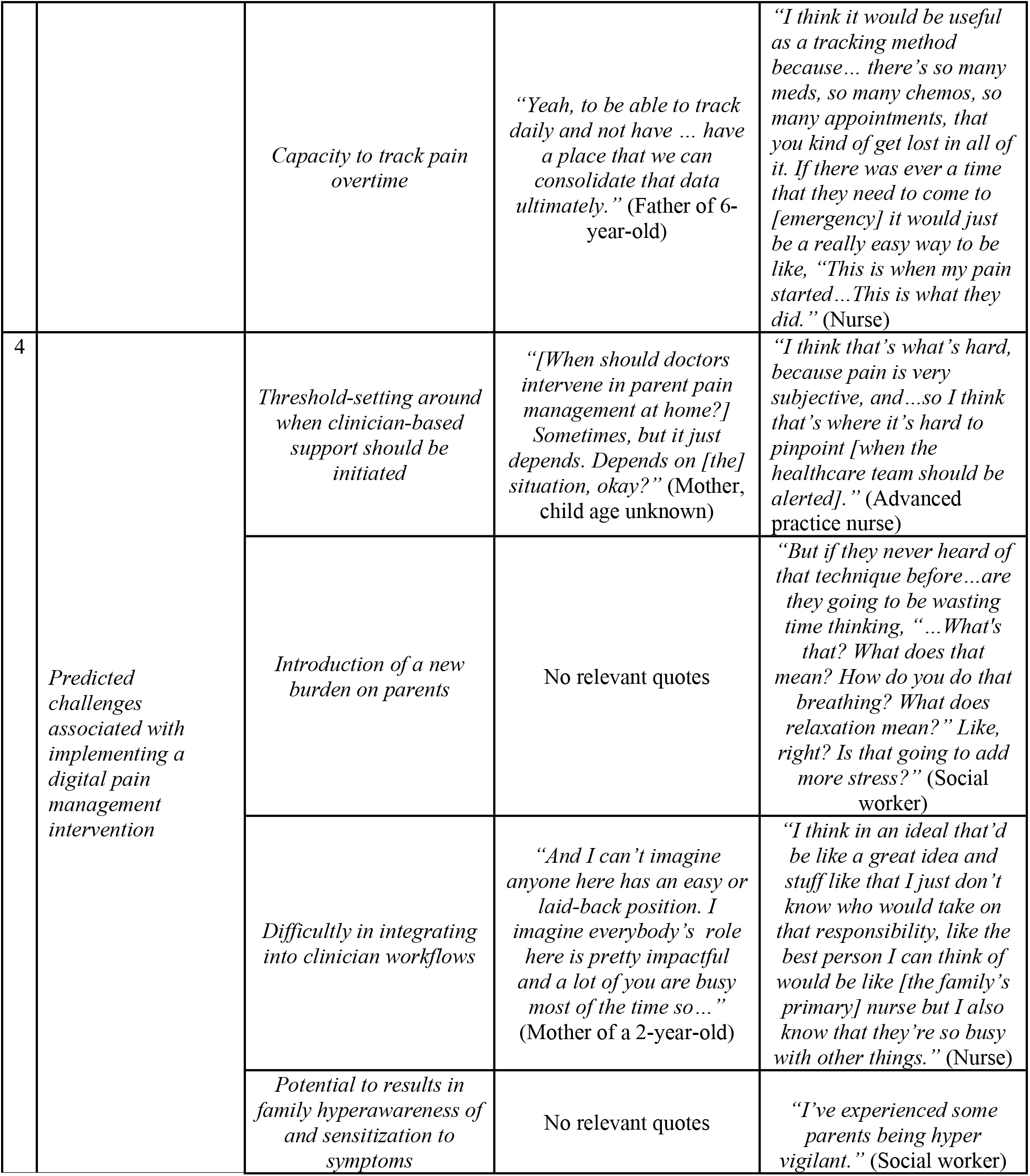
Thematic analysis framework

### The need for and value of a digital cancer pain management intervention

Both parents and caregivers considered a pain management app as a useful, safe, and convenient way to empower caregivers to effectively manage their young child’s pain in the home environment, especially for parents of children newly diagnosed with cancer. Several current deficiencies with at-home pain management for young children were also discussed. Informational and procedural support for parents was needed and valued. Both groups noted that parents already possessed a strong suite of child pain management skills, but a means to support the implementation of this knowledge “would be helpful” (Mother of a 6-year-old). Clinicians described current clinical practices of learning about young children’s cancer pain as not comprehensive and often reliant on reviews of at-home medication use rather than lived experiences of pain. Procedural support to allow parents “to track when [their child is] having the pain and how long it’s been” (Nurse) would provide in-depth information to healthcare teams and facilitate communication between parents and clinicians. Parents and clinicians also perceived pain to be better managed in hospital and described a need for connection to the interdisciplinary team and associated diverse treatment strategies at home. Parents also alluded to the often overwhelming task of managing pain at home stating, “You panic sometimes when your child is sick. You’re like, “What’s going on? How much [pain medication] do I give? What do I do?” (Mother of a 4-year-old). The numerous other direct and indirect cancer care tasks for which parents assumed responsibility—such as medication administration and appointment tracking—amplified this strain and a tool that could offer emotional support to parents was considered advantageous.

### Recommendations pertaining to pediatric digital health interventions in general

Parents and clinicians made recommendations that might be applied broadly to most parent-led digital health interventions, beyond just apps targeting pediatric cancer pain. These features were suggested to enhance intervention usability, credibility, and resultant effectiveness. Participants particularly noted the need to embed accessibility features in intervention design, emphasizing that these “app[s] should be made as easy as possible so that anybody can use” (Mother of a 4-year-old). To improve intervention access for parents, participants recommended that health apps be available in multiple languages, primarily use lay terms for caregivers “who [are] not in the medical field to understand” (Father of a 6-year-old) and include a text-to-audio feature and supplemental information that users could review as needed. Participants also recommended integrating a calendar feature to act as a centralized hub with scheduled medication and medical appointment reminders, and allow for child symptom tracking. Many participants also suggested adding gamified elements such as avatars and pictorial representations. Such gamification was considered to encourage family-oriented digital care by “making it kid friendly” (Stepmother of a 10-year-old) and would encourage child engagement in app-based care alongside their caregiver. Specific recommendations related to the use of avatars were to include features reflective of the parent and child (e.g., diverse skin tones, range of heights and weights, accessibility features such as glasses and prosthetics).

Various features that would enhance the credibility of a digital interventions were also highlighted. Clinician integration within digital health interventions was considered important. Considering this integration, participants recommended a software-embedded “threshold for [symptom] follow-up” (Nurse), that if surpassed, would result in notification to the healthcare team. Such notifications were suggested to ensure child safety and to build technologies that enhance, as opposed to replace, care delivered by clinicians. This sharing of app-generated health reports with clinicians was also considered an important means to support symptom management by “sort of opening a door to a conversation that maybe [parents and clinicians] wouldn’t have had” (Oncologist). To enhance credibility, participants also strongly recommended that digital interventions be based on sound and up-to-date scientific evidence-based and care recommendations be personalized to users, including by implementing software algorithm-based personalization. Finally, clear descriptions of the limits of clinical support that could be provided by a digital health intervention should be embedded. Participants highlighted that such disclaimers would build trust with the intervention by emphasizing when care outside of that provided by the app should be sought.

### Recommendations pertaining to parent-led digital cancer pain management specifically

Participants suggested four categories of priority features for effective caregiver-led digital pediatric cancer pain management. First, participants suggested the inclusion of a feature enabling the storage of a child’s pain history, medical history, current medications, allergies, social history, and family history. This information could be updated by both parents and clinicians and would support the personalization of pain management advice for a child.

Second, suggestions were made related to app-based pain assessments. Both parents and clinicians strongly recommended multi-dimensional pain assessment tools embedded in the app to measure pain as a sensory phenomenon (e.g., pain frequency, duration, severity, and cause) and consider the impact of pain on a child’s affective state and a child’s activities of daily living, including ability to take medications. App-embedded pain assessments should be valid and implemented “in a moment when [pain is] actually happening” (Nurse) for the sake of accuracy. Assessments should also be appropriate to the age, developmental stage, and abilities of the younger child with cancer and should be repeated once a pain management intervention was implemented. Therefore, parent proxy assessments should be embedded for children unable to self-report their pain. Parents also suggested building in capacity to record other aspects of their child’s physical status that may be related to pain (e.g., bowel movement patterns, nausea and vomiting). Caregivers also discussed the utility of an application that provides developmentally appropriate management advice, specifically citing a need for “proper information for that age group” (Father of a 5-year-old), whenever pain is reported. Parents felt confident in their abilities to understand and implement app-based pain management instructions, noting familiarity with many mobile apps, including health apps, they currently use.

Multi-modal physical, psychological, and pharmacological cancer pain management advice was recommended to be provided to parents. App-integrated physical advice included “gentle exercise” (Physical therapist) and stretching. Recommended psychological advice was distraction including “playing” (Father of a 6-year-old), relaxation, and massage, and advice to emotionally support parents in their role as pain caregivers was also encouraged. Parents suggested that the app provide medication advice so long as the information presented was evidenced-based and specific to the context of pediatric cancer. Embedding information on how to manage medication side effects was also suggested.

Fourth, both parents and clinicians expressed a need for a digital intervention to support the tracking of multidimensional child pain assessment data overtime and provide visual trends of the data, with the intention of “pick[ing] up patterns” (Oncologist) and supporting family-clinician conversations on pain. Within the app, parents and clinicians also recommended tracking which pain management strategies were used, and the effectiveness of each strategy.

### Predicted challenges associated with implementing a digital pain management intervention

Although participants cited digital health intervention aspects that were considered beneficial, predicted challenges were also highlighted. Most prominently, participants discussed potential difficulty in setting a software-embedded pain threshold beyond which healthcare provider support would be initiated. Participants specifically discussed the subjective nature of pain and a resultant need to individualize pain thresholds to each child and their family context. Clinicians discussed the potential for the app to add burden for parents who are already strained with caregiving tasks for their child; however, several parents considered use of the app, including once or twice a day, to “be feasible” (Father of a 6-year-old). Clinicians further cited challenges related to how to successfully integrate app-based data monitoring and pain support into their daily workflows. In both cases, participants highlighted that the demonstrated positive impact of the intervention on child pain outcomes must outweigh any burden of use. Participants also worried that frequent pain monitoring facilitated by the app may lead parents and children to become acutely aware and subsequently hypersensitized to pain.

## DISCUSSION

This study investigated the perspectives of parents of young children with cancer and pediatric cancer care clinicians as they pertain to the design and delivery of digital health interventions targeting childrens’ pain. Woven throughout the recommendations were the need for and value of such pain management interventions, especially in light of the limited at-home family-led pain management strategies and the often overwhelming nature of managing pain independently in care settings other than the hospital. Parents and clinicians recommended ways to improve the usability and credibility of such interventions, including the integration of accessibility features and ensuring high-quality pain management content is delivered. Participants recommended needed features for a digital pediatric cancer pain management intervention including embedded multidimensional assessments of pain, multi-modal pain management support, and pain tracking over time. Critical to such assessments was the inclusion of detailed child and family information to support the personalization of provided pain support. Lastly, stakeholders noted perceived challenges associated with such interventions that should be addressed to enhance utility.

Existing literature highlights the critical importance of involving users in the development and evaluation of apps for specific health conditions, particularly noting the positive impact on intervention use and effectiveness in achieving an intended health goal (22). Meaningful stakeholder engagement also enables understanding of the key design features that will facilitate successful digital interactions, and those features that may limit use (e.g., poor integration with daily workflows) (22–24).

Our study participants reinforced the need for demonstrated usability and credibility within digital health interventions for children, including those targeting young children’s cancer pain. Recommendations for accessible language features, the use of lay terms and supplemental health information, and the use of gamification to engage younger users are supported by the existing literature (25,26). Research shows that language barriers in healthcare lead to miscommunication between medical professionals and patients. The implementation of language accessibility features improves the quality of healthcare delivery and patient safety (27). App gamification has also been shown to improve the user experience, accessibility, app appeal, user experience, while also having a positive impact on well-being and health outcomes (28).

Our results also show the critical importance of using evidence-based information and clinician involvement in providing app-based health advice to users. These needed app features were endorsed by parents and reflect the findings of a recent systematic review that identified standards for mobile health applications, including that apps should name content authors and their professional qualifications and utilize scientific evidence as the basis for quality content (29). Further, participants in our study recommended the implementation of disclaimers in digital health intervention, again aligning with current health app guidance to disclose possible risks to users and include warnings that apps are not intended to replace health professional care (29).

The need to embed a comprehensive multi-dimensional pain assessment tool and multi-modal integrative pain management advice in the app reflects current considerations for the management of cancer-related pain. This recommendation reflects a clinical need to implement a biopsychosocial approach to pain assessment and management, including treating childhood cancer pain with evidence-based pharmacological, psychological, physical, and complementary and alternative medicine techniques (1). Consideration of the time to complete comprehensive assessments is also needed as previous research in pediatric cancer pain apps has shown lengthy questionnaires to be a drawback to app use (13).

Predicted challenges associated with digital health interventions identified in our study include the potential for the app to burden parents and clinicians. Digital burden has been described previously and studies of high-burden apps often experience high participant attrition rooted in the fundamental challenges of keeping participants engaged in intervention use (30). Difficulties in setting app-based pain thresholds for the initiation of clinician-driven pain management support require the application of a personalized approach to pain care (31) and a need to further tailor co-designed digital interventions to the needs of unique users and contexts. (32)

Strengths of our study include the integration of multiple stakeholders’ perspectives and our collection of data from two pediatric cancer care centers. Limitations include potential social desirability response bias whereby parents and clinicians may have withheld negative reports about the app concept during interviews with our team and we did obtain the necessary purposive sample. However, participants were informed that all feedback would be valued equally. Additionally, the mode by which we conducted interviews was not standardized due to the COVID-19 pandemic and we instead held interviews via telephone, online, and in-person. However, research shows good comparability between the content and depth of interviews conducted in-person and otherwise (33,34). Finally, we did not conduct participant checking of either the transcriptions or final thematic analysis.

This study provides recommendations from key stakeholders on the design and delivery of digital pediatric cancer pain interventions, which can be readily used by parents, clinicians, engineers, and researchers participating in intervention development. Further, several recommendations gleaned from this process are directly applicable to the design of many pediatric digital health interventions—including those beyond childhood cancer pain. We recommend that future investigators engaged in developing health apps implement a co-design approach within their work. Such participation can reveal critical requirements for intervention design that result in relevance of app content and enhanced clinical effectiveness. The recommendations gathered in the present study will become the design principles of our future childhood cancer pain digital research.

## MATERIALS AND METHODS

Our reporting is in accordance with the Consolidated Criteria for Reporting Qualitative Research (COREQ) (15) and Guidance for Reporting Involvement of Patients and the Public—Short Form (GRIPP2-SF) (16).

### Study Approach, Setting and Participants

We used an inductive, qualitative descriptive approach for healthcare research (17), consistent with our goal to understand stakeholder’s pain app perceptions and requirements. We recruited participants from the Hospital for Sick Children (SickKids) in Toronto, Ontario, Canada and the Children’s Hospital of Orange County (CHOC) in Orange, California, United States. We enrolled English-speaking parents who were the primary caregiver of a child (2-11 years) receiving treatment for any cancer diagnosis and who spent at least 25% of their cancer-care treatment time outside of the hospital and who had pain of any intensity in the preceding week. Child pain was determined by caregiver proxy-report. English-speaking multidisciplinary clinicians were included if they worked within the hematology/oncology program at either hospital and provided direct pain-related care to children spending at least 25% of their time during cancer treatment at home. We employed a purposive maximum variation sampling strategy with the aim of including parents who varied in age, sex, ethnicity, and their child’s diagnosis, and clinicians who varied by healthcare profession.

### Data Collection

Following ethics board approval at both sites, we obtained informed consent from participants. Participants were asked to complete demographic questionnaires and parents completed a Parental Pain Expression Perceptions (PPEP). The PPEP is a valid and reliable 9-item Likert-type scale-based questionnaire that assesses parents’ knowledge and attitudes about pain expression in children, where higher scores represent greater misconceptions about pain expression (18).

Individual in-hospital, telephone, or online face-to-face semi-structured interviews were conducted by trained research team members (AC, HGP, LBT) with no previous relationship to participants. Our interview guide was based on the currently developed Pain Squad+ pain management app for adolescents with cancer (19) and a study of salient home-based pediatric cancer care issues (20). We audio-recorded all interviews and handwritten field-notes were taken. We conducted interviews concurrent with our analyses until themes relevant to the study’s aim reached saturation.

### Data Analysis

Interview audio-recordings were transcribed into electronic documents and uploaded to NVivo software version 11.4.0. Using an inductive approach, transcripts were coded with reference to field-notes by four independent research assistants (SS, KH, TM, and MZ). Following the method of Braun and Clarke (21), we read through the dataset multiple times and discussed its features as a group before creating several coding categories. Coding proceeded using a statement-by-statement approach and codes were organized into themes and subthemes. The creation of themes was an iterative process where themes were continuously reviewed and compared against the narrative exemplars until a final thematic framework was established. The research team (SS, KH, TM, and LJ) met multiple times to ensure themes were accurately reflective of participant narratives, and consensus building exercises were used to refine themes and subthemes where needed.

## Data Availability

Data are available from Dr. Jibb for researchers who meet the criteria for access to confidential data.

## Acknowledgements

We would like to thank our parent and clinician participants for kindly taking the time to share their experiences and perceptions related to the design and development of a future app. We would also like to thank Alyssandra Chee-a-tow for her invaluable assistance with interviewing.

## Abbreviations

CHOC: Children’s Hospital of Orange County
PPEP: Parental Pain Expression Perceptions
SickKids: The Hospital for Sick Children

